# Public Preferences for Government Response Policies on Outbreak Control

**DOI:** 10.1101/2020.09.04.20187906

**Authors:** Semra Ozdemir, Si Ning Germaine Tan, Isha Chaudhry, Chetna Maholtra, Eric Andrew Finkelstein

## Abstract

**Objectives:** To assess the extent to which public support for outbreak containment policies varies with respect to the severity of an infectious disease outbreak.

**Methods:** A web-enabled survey was administered to 1,017 residents of Singapore during the COVID-19 pandemic, and was quota-sampled based on age, gender and ethnicity. A fractional-factorial design was used to create hypothetical outbreak vignettes characterised by morbidity and fatality rates, and local and global spread of an infectious disease. Each respondent was asked to indicate which response policies (among 5 policies restricting local movement and 4 border control policies) they would support in 5 randomly-assigned vignettes. Binomial logistic regressions were used to predict the probabilities of support as a function of outbreak attributes, personal characteristics and perceived policy effectiveness.

**Results:** Likelihood of support varied across government response policies; however, was generally higher for border control policies compared to internal policies. The fatality rate was the most important factor for internal policies while the degree of global spread was the most important for border control policies. In general, individuals who were less healthy, had higher income and were older were more likely to support these policies. Perceived effectiveness of a policy was a consistent and positive predictor of public support.

**Conclusions:** Our findings suggest that campaigns to promote public support should be designed specifically to each policy and tailored to different segments of the population. They should also be adapted based on the evolving conditions of the outbreak in order to receive continued public support.

## 1. Introduction

In deploying response policies to contain an infectious disease outbreak, governments have to balance controlling its spread against restricting personal liberty [1, 2] and adverse economic outcomes [3]. Governments have to adapt and adjust policies at different phases of an outbreak, and in most cases, with limited and evolving information. In doing so, governments are concerned about the public opinions of their constituencies especially when public support is paramount to ensuring successful implementation of these policies [2, 4-6]. Only a handful of studies have investigated public support for government containment policies for a pandemic and the factors that influence acceptance [6-9]. However, they did not investigate public support for more restrictive policies, such as shutting down public transportation or initiating a lockdown, which have been implemented in multiple countries during the COVID-19 pandemic. Additionally, none but one [8] accounted how public support for government measures varied with respect to pandemic characteristics, and only one study was actually conducted during a pandemic [9].

The main objective of this study was to assess the extent to which public support for government response measures varied with respect to the severity of an infectious disease outbreak. The survey was administered to an adult sample of Singapore’s general population during the initial stages of the COVID-19 pandemic. This provided the unique opportunity to capture public opinion in a period of potential physical, psychological and financial turmoil of an outbreak-turned-pandemic [10-13].

An individual’s support of a specific public health policy likely depends on how they perceive its benefits and costs to their household, community as well as to society [14]. Our hypothesis is that public support for a specific outbreak control policy will be higher as per rates of morbidity and fatality within the country and its spread locally and across the world. We also hypothesize that fatality rate will be most important factor for internal policies, while global spread will be the most important factor for border control policies.

We also investigated how likelihood of support varies with personal characteristics. An earlier study indicates that individuals with lower income are less likely to support government response policies [6], possibly because they are more likely to experience loss of income due to these policies [15]. All else equal, older and less healthy individuals are more likely to support government policies as they are more vulnerable and adversely affected by infectious diseases than younger and healthier individuals [16, 17]. Since income and health effects are expected to work in opposite directions, we investigated which is more dominant. Literature also showed that measures perceived to be more effective (e.g. frequent handwashing and mask-wearing) were practiced more compared to those that were not perceived to be so (e.g. working from home) [18, 19]. Hence, we also investigated whether individuals who perceive specific policies to be effective are more likely to support them.

## 2. Methods

### 2.1 Study Sample

A web-enabled survey was administered to a panel of individuals from the general population in Singapore, recruited through a market research company. Participants had to be aged 21-years old and above (the legal age in Singapore) and residents of Singapore. We quota-sampled based on gender, age and ethnicity to ensure national representation based on these characteristics. All respondents provided written informed consent and received incentives in the form of a points-based reward awarded by the survey company. The study was exempted from review by the Institutional Review Board of National University of Singapore (Application Reference Number: S-20-085). The survey was fielded between March 31^st^ and April 14^th^ 2020 during the COVID-19 pandemic. When the survey was launched on March 31^st^, there were 926 confirmed cases of COVID-19 and 3 COVID-related deaths in the country [20].

### 2.3 Survey Development

Respondents were shown 5 vignettes describing the early days of an outbreak. Each vignette was characterized using five attributes (Supplementary Material A). Two of the attributes described the degree of spread of the disease locally and globally, respectively: “Total number of confirmed cases in Singapore since the outbreak started” and “Number of countries with rapidly increasing number of cases”. To express the trend of the local disease spread, we used: “Number of new cases in Singapore within the last 2 weeks”. Two other attributes described the morbidity and fatality of the disease: “Number of cases admitted to intensive care unit (ICU) but did not end in death in Singapore” and “Number of infection-related deaths in Singapore”. Information on the selection of attribute levels can be found in Supplementary Material B. Due to restrictions on recruitment for research studies because of COVID-19, we tested the survey instrument with 9 staff members (not in the research team) using a think-aloud protocol.

The experimental design for the vignettes was created based on fractional-factorial design in SAS and included 50 vignettes. The experimental design allowed for interaction effects between the total number of confirmed cases and 1) the number of new cases in the last 2 weeks; 2) the number of ICU cases; and 3) the number of deaths. We grouped the vignettes into 10 blocks of 5 to reduce the respondent’s cognitive burden. Each respondent was randomly assigned to one block.

Respondents were introduced to the attributes as factors that the government takes into account when responding to emerging infectious disease outbreaks. It was emphasised that this was not pertaining to COVID-19, but any potential outbreak that could occur in the future. For each vignette, respondents were asked to select which policies the government should implement. The policies included 5 internal policies which restrict movement within the country, and 4 border control policies. Respondents could select as many policies as they preferred or could select none. Figure 1 presents an example vignette.

**Figure 1.**
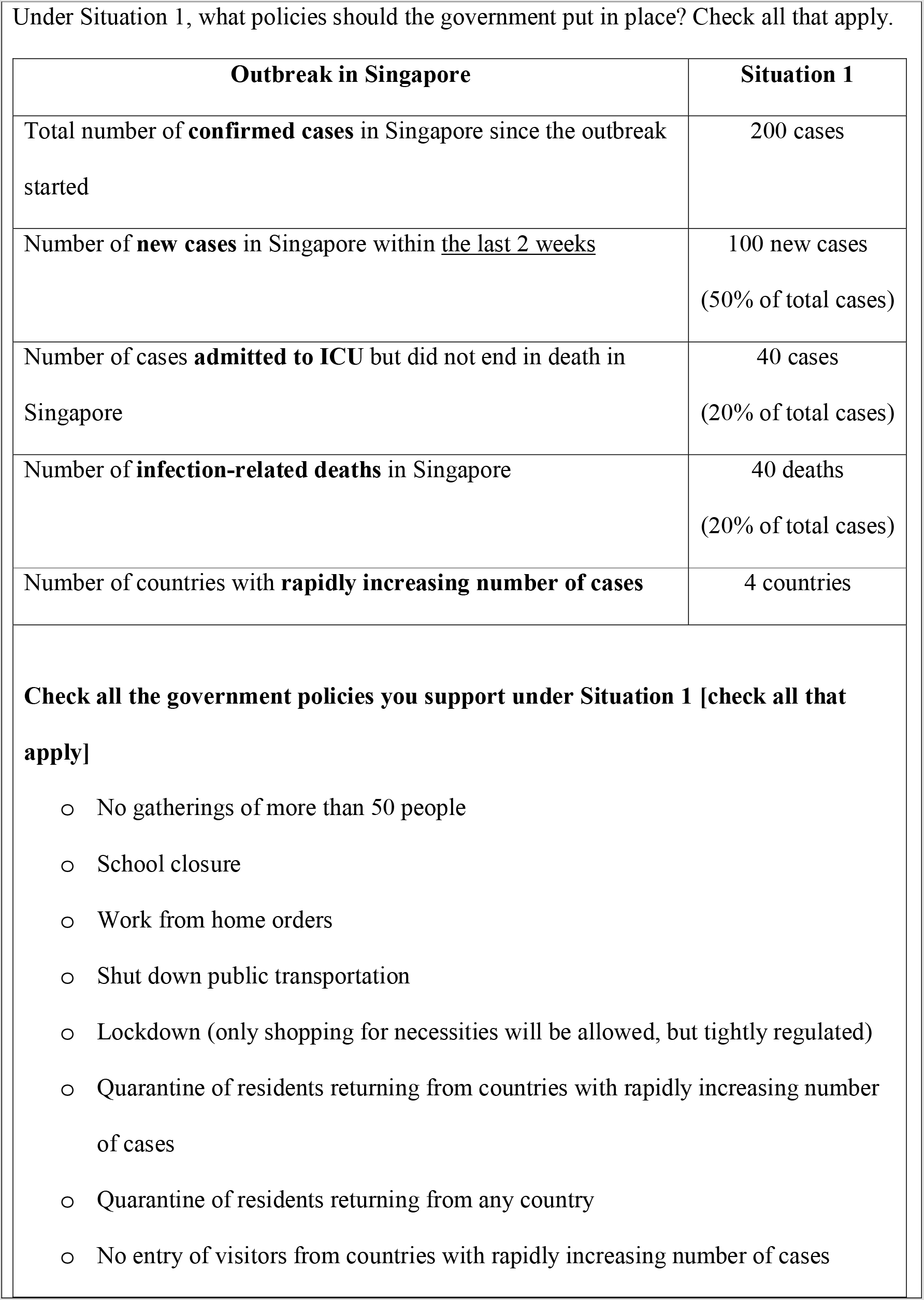
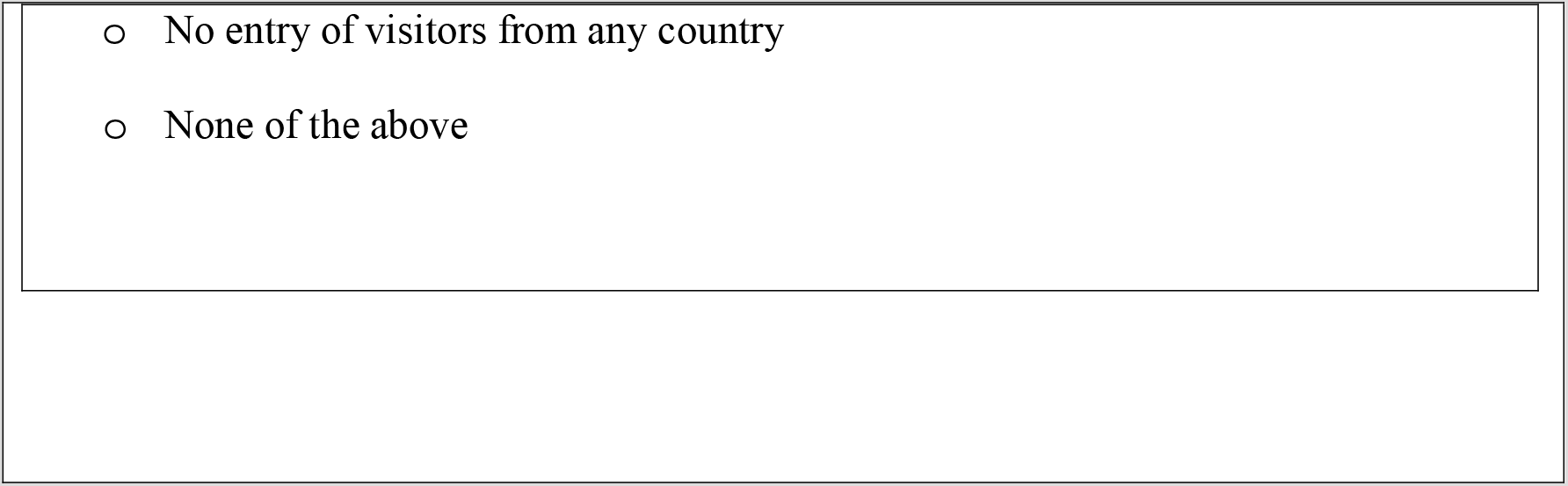
Example vignette.

### 2.4 Statistical Analysis

We used binomial logistic regressions to estimate the probability of choosing a policy where ‘1’ indicated support for the policy and ‘0’ indicated no support. Since each respondent was shown multiple vignettes, to account for the selections by the same respondent, the regressions were estimated with clustered standard errors where the clustering was based on respondent identification.

The independent variables included 5 outbreak-related attributes described above and personal characteristics. The main effects and interaction effects on outbreak attributes were entered into the model as continuous variables. Based on our research questions, the perceived effectiveness of a policy (“very effective”, vs “somewhat effective” or “not effective”), age, self-reported overall current health status (poor or fair vs good, very good or excellent), and income were included as predictors. We used housing type as a proxy for income categorisation since it was less likely to be affected by the short-term effects of COVID-19. As land is limited and housing is expensive in Singapore, majority of the residents live in flats built by the Housing Development Board (HDB) on government-owned land which are colloquially referred to as HDBs. Based on the monthly incomes reported for each housing type [21], we categorised 1 – 3 room HDBs as lower income, 4 – 5 room HDBs as middle income, and private condominiums/apartments and landed houses as high income. We also investigated the interaction effects between health and income dummy variables to investigate whether concerns related to income moderate the effect of health concerns (and visa verse). Although we did not have a specific hypothesis related to ethnicity, we included a dummy variable for the Chinese ethnicity (vs other racial/ethnic groups) as a control variable since Singapore is home to multiple ethnic groups.

We first presented the percentage of times each policy was selected by the respondents (averaged over all vignettes). Since the interpretation of coefficients is complicated when a logistic model has both main and interaction effects, we calculated the average marginal effects for the outbreak attributes to identify the relative importance of each attribute. The average marginal effects provide the average change in probability for the sampled population when an attribute increases by one unit. We also predicted the average probability of support (i.e. choosing a policy) for each policy under 4 scenarios (2 that are less severe and 2 that are more severe) to show how the likelihood of support changes based on the varying severity of the outbreak. To assess the effects of health and income on policy support, we presented the average predicted probabilities for different subpopulations (e.g. less healthy and low-income groups). Lastly, we presented the average marginal effects of age and perceived effectiveness of a policy on the likelihood of support. Stata 16.1 was used to perform the analyses.

## 3. Results

### 3.1 Respondent Characteristics

1,017 respondents completed the survey. Over half (51%) of the respondents were female (Table 1). The majority (80.1%) were Chinese with a median age of 40 years. There was a relatively uniform distribution of age groups, with 35.2% aged between 21 to 35 years old, 35.4% aged between 36 to 50 years old, and 29.4% aged above 50 years old. Overall, the sample was representative of the national population in terms of gender (51% females) and age (median: 41.1 years), but over-represented Chinese ethnicity (74.3% of the population) [22]. More than half were married (55.7%) and over half the sample had a university degree (58.1%). Most respondents (67.4%) were employed full-time while 15% were employed part-time or self-employed. Based on housing type, 19.5% was categorized as lower income, 59.3% as middle income and 21.2% as higher income. The distribution of housing type in our sample is similar to national statistics on housing [23]. About 17% reported having fair or poor health status at the time of survey.

**Table 1.** Demographic characteristics, N = 1017.

| **Characteristic** | **N (%)** |
| --- | --- |
| Female | 519 (51.0) |
| Age, Median (IQR) | 40 (32, 52) |
| Ethnicity |  |
| Chinese | 815 (80.1) |
| Malay | 91 (9.0) |
| Indian + Others | 111 (10.9) |
| Married | 567 (55.7) |
| Education |  |
| Below secondary | 139 (13.7) |
| Vocational/Diploma | 287 (28.2) |
| University or above | 591 (58.1) |
| Employment |  |
| Full-time | 686 (67.4) |
| Part-time/Self-employed | 153 (15.0) |
| Others | 178 (17.5) |
| Housing and income |  |
| 1 – 3 bedroom public housing (low-income) | 198 (19.5) |
| 4 – 5 bedroom public housing (middle-income) | 603 (59.3) |
| Private housing | 216 (21.2) |
| Current health status |  |
| Excellent/very good | 383 (37.7) |

| Characteristic | N (%) |
| --- | --- |
| Good | 459 (45.1) |
| Fair/poor | 175 (17.2) |

We observed variation in perceived effectiveness of each policy reported by the respondents (Table 2). The policies that received the highest proportions of the ‘very effective’ rating were ‘no entry of visitors from countries with rapidly increasing number of cases’ (henceforth referred to as banning visitors from selected countries) (75.7%) and ‘quarantine of residents returning from countries with rapidly increasing number of cases’ (henceforth referred to as quarantining residents from selected countries) (70.2%). The policies that received the lowest proportions of ‘very effective’ rating were shutting down public transportation (32.1%) and ‘no gatherings of more than 50 people’ (henceforth referred to as banning large gatherings) (39.4%).

**Table 2.** Perceived effectiveness of a policy and percentage of times a policy was selected, N = 1017.

| **Policy** | **Perceived effectiveness of policy**<br>**(very effective)** | **Percentage of times a policy was selected** |
| --- | --- | --- |
| No gatherings of more than 50 people | 39.4% | 74.3% |
| School closure | 41.0% | 55.7% |
| Work from home orders | 54.9% | 71.4% |
| Shut down public transportation | 32.1% | 30.4% |
| Lockdown (only shopping for necessities will be allowed, but tightly regulated) | 52.4% | 45.1% |
| Quarantine of residents returning from countries with rapidly increasing number of cases (i.e. restricted countries) | 70.2% | 80.7% |
| Quarantine of residents returning from any country | 67.2% | 75.6% |
| No entry of visitors from countries with rapidly increasing number of cases (i.e. restricted countries) | 75.7% | 78.6% |
| No entry of visitors from any country | 68.2% | 64.9% |

### 3.2 Vignette Findings

Table 2 presents the percentage of times each policy was selected. The policies that were selected the most were quarantining residents from selected countries (80.7%) and banning visitors from selected countries (78.6%). The policy that was selected the least was shutting down public transportation (30.4%), followed by lockdown (45.1%).

Results from the logistic regressions are presented in the Supplementary Material C. Table 3 shows the average marginal effects for all the policies. As we hypothesized, the fatality attribute had the highest average marginal effects for most internal policies with the largest effect observed for lockdown (0.0054, p-value<0.001), followed by school closure (0.0048, p-value<0.001); however, it was not significant for banning large gatherings (0.0002, p-value = 0.82). The number of countries with rapidly increasing number of cases had the highest average marginal effects for 3 (out of 4) border control policies with the largest effect observed for ‘no entry of visitors from any country’ (henceforth known as banning all visitors) (0.0063, p-value<0.001). None of the outbreak-related attributes were significant for quarantining residents from selected countries.

**Table 3.**
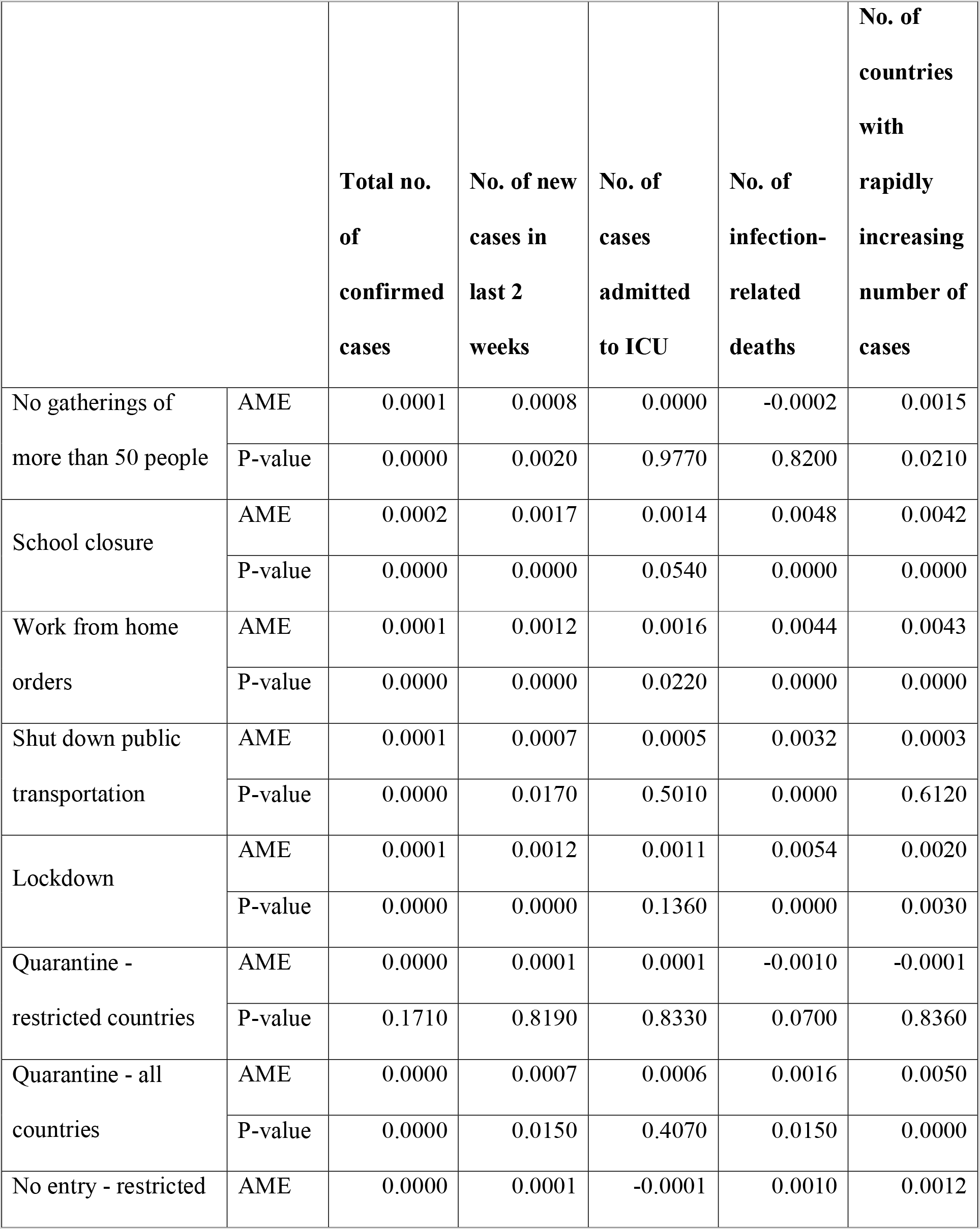
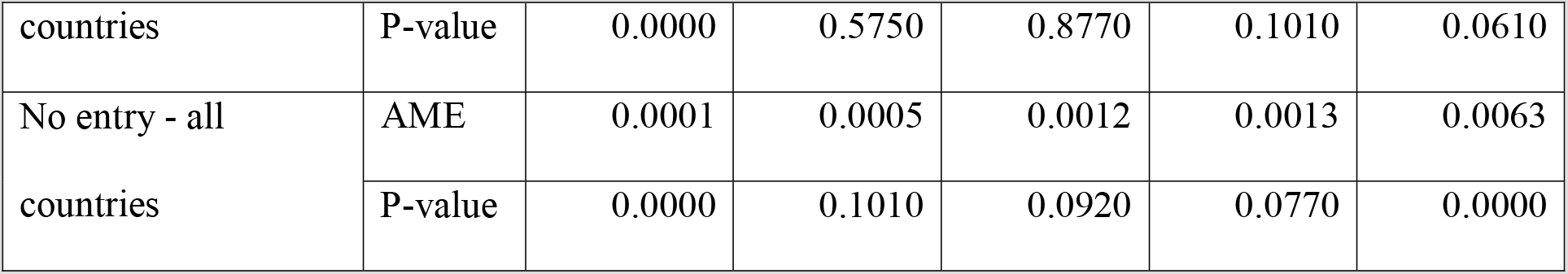
Average marginal effects (AME) for outbreak attributes.

The average predicted probabilities of support (including 95% confidence interval (CI)) for each policy in four different scenarios are presented in Figure 2 (the full list of probabilities, CIs and p-values are presented in Supplementary Material D. Scenarios 1 and 4 were the least and the most severe scenarios respectively, and therefore they should receive the lowest and highest probabilities of support accordingly for each policy. The number of countries with rapidly increasing cases was kept constant (at 4) across all the scenarios to highlight the variation in predicted probabilities due to country-specific factors only.

**Figure 2.**
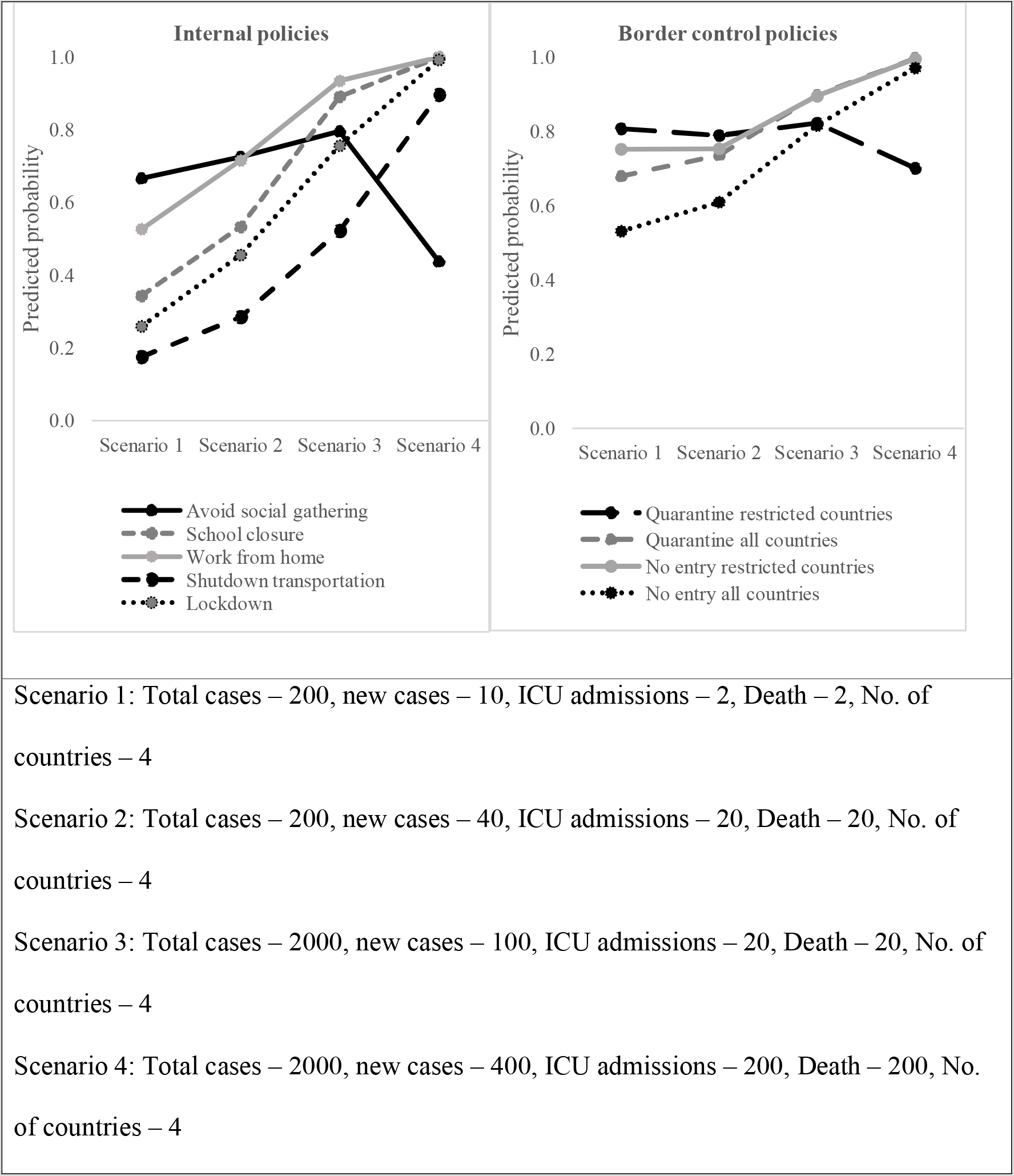
Average predicted probability of support for a policy for varying levels of outbreak severity.

Among the internal policies, the probability of support for the least severe scenario (Scenario 1) was the highest for banning large gatherings (0.67 [0.63, 0.70]), followed by work from home orders (0.53 [0.49, 0.57]), and was the lowest for shutting down public transportation (0.17 [0.15, 0.20]) (Figure 2). The probability of support was greater in scenarios with more severe outbreak scenarios for all policies, except for banning large gatherings. The steepest increase in support from Scenario 1 to Scenario 4 was for lockdown (from 0.26 [0.23, 0.29] to 0.99 [0.96, 1.02]), followed by shutting down public transportation (from 0.17 [0.15, 0.20] to 0.90 [0.50, 1.29]).

Among the border control policies, the probability of support in Scenario 1 was highest for quarantining residents from selected countries (0.81 [0.78, 0.84]), and lowest for banning all visitors (0.53 [0.49, 0.57]). The probabilities for Scenario 1 for border control policies were generally higher than the probabilities for internal policies. For quarantining residents from selected countries, the probabilities remained relatively stable for different levels of outbreak severity as the outbreak attributes were not significant. Policy on banning all visitors had the steepest increase in probability from Scenario 1 to Scenario 4 (from 0.53 [0.49, 0.57] to 0.97 [0.84, 1.10]).

Among respondent characteristics, lower-income dummy variable was significant and negative for quarantining residents from selected countries and banning visitors from selected countries (p-value<0.05) while middle-income dummy variable was not significant for any of the policies (p-value>0.10). Less-healthy dummy variable was significant and positive for all 4 border control policies (p-value<0.10) (Supplementary Material C). Interaction effects between health and income dummy variables were not significant for any of the policies (results not shown), thus, the final models included only main-effects of health and income.

When we investigated likelihood of support for population sub-groups, we found that the average predicted probabilities of policy support were higher (as expected) for the less healthy compared to the healthy in each income group, and the differences were statistically significant for all border control policies (p-value<0.10). We then investigated the income effect for less-healthy and healthy individuals by comparing lower- and middle-income groups to high-income group. We found that the average predicted probabilities were lower (as expected) for the lower-income group compared to the high-income group, and the differences were significant for both less healthy and healthy individuals for only 2 border control policies (p-value<0.10). The average predicted probabilities were not significantly different between the middle-income group compared to the high-income group (for both less healthy and healthy individuals) for any policy (p-value>0.10). (Table 4 for differences in average predicted probabilities; Supplementary Material E for the full list).

**Table 4.**
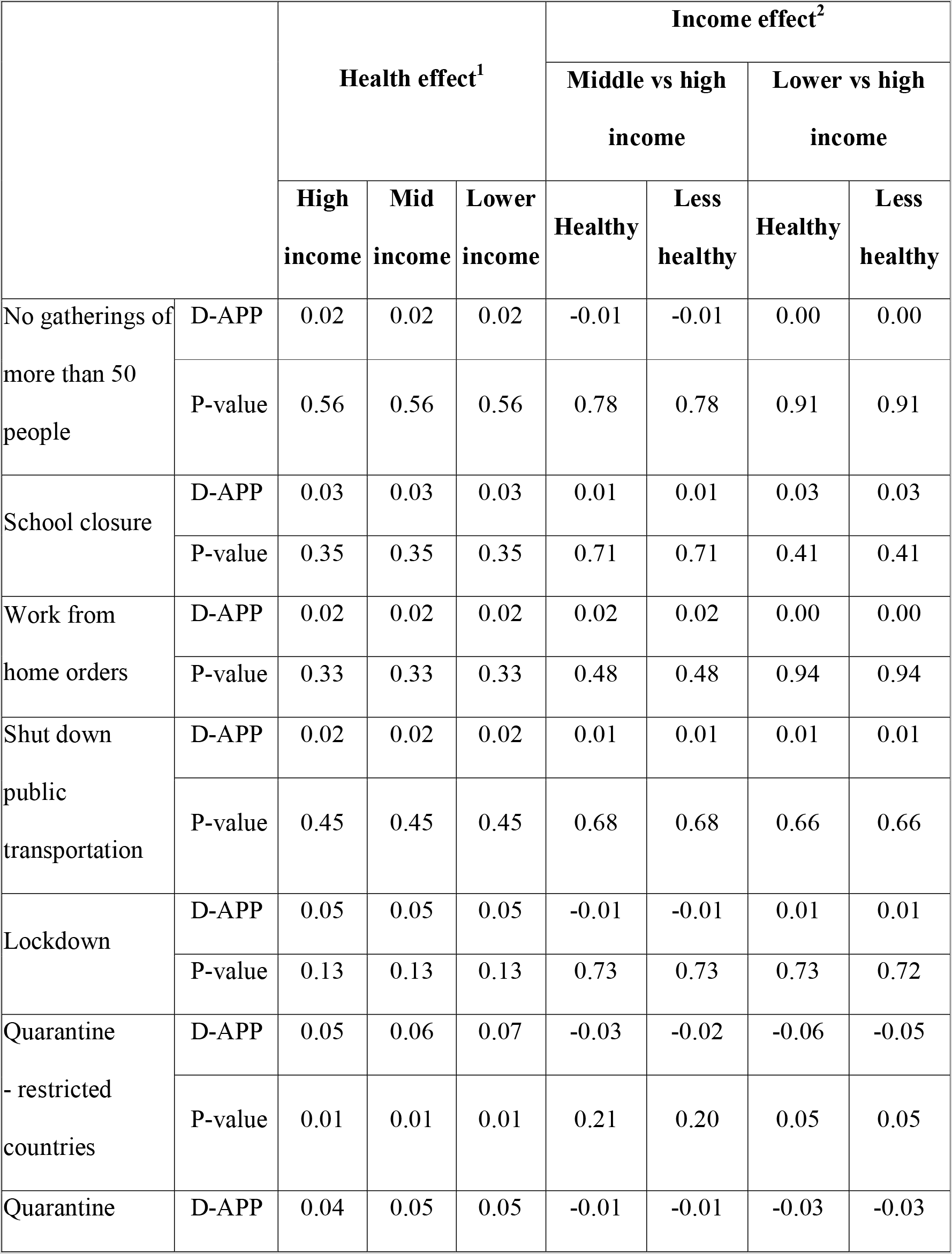
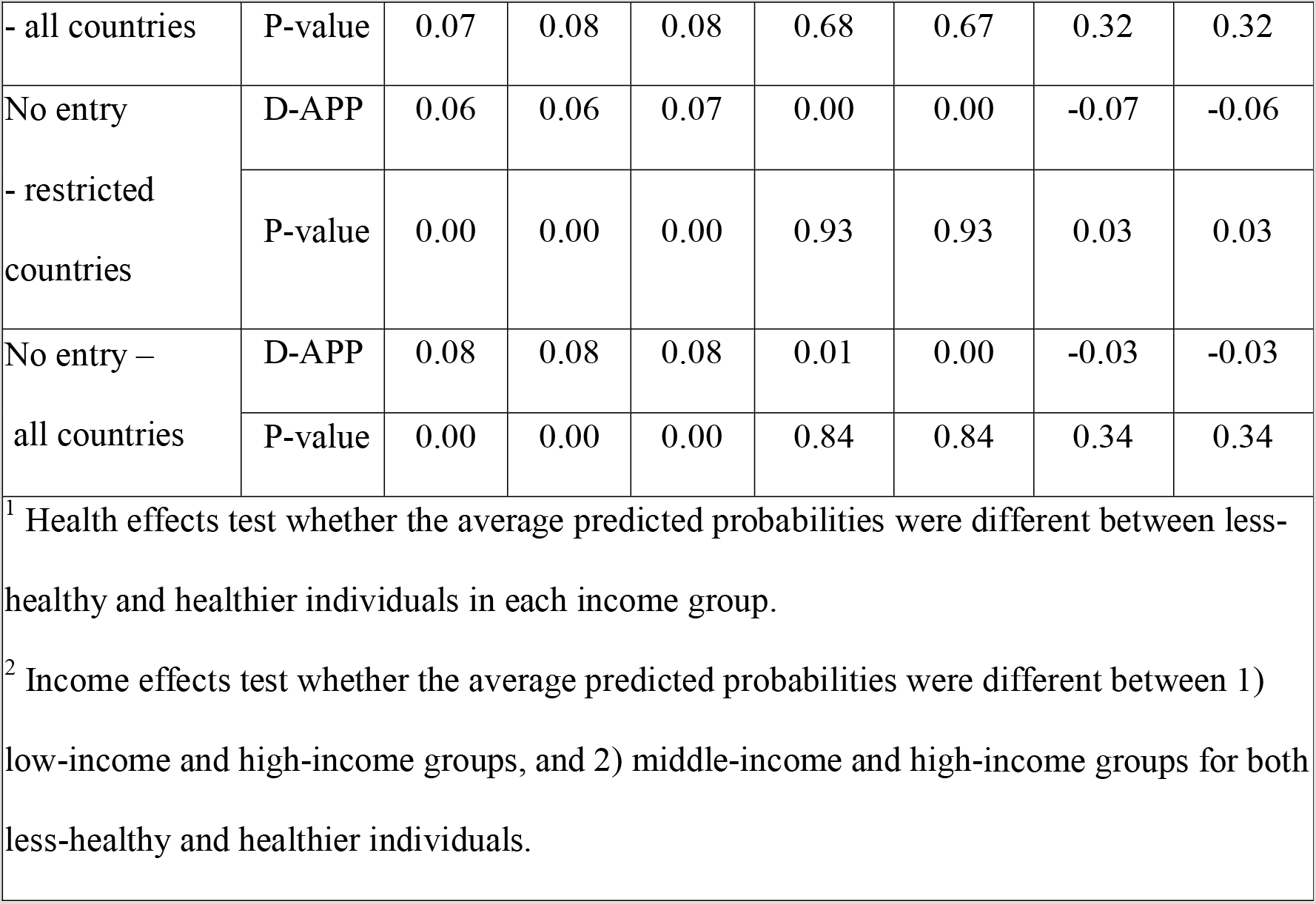
Differences in average predicted probabilities (D-APP) for sub-groups based on health and income.

Our findings show that older (vs younger) patients were more likely to support banning large gatherings, working from home and all border control policies while they were less likely to support shutting down public transportation (p-value<0.10 for all). The average marginal effects of age were largest for banning large gatherings (0.003, p-value<0.01) and banning visitors from selected countries (0.003, p-value<0.01) (Supplementary Material F). The probabilities of support were significantly higher for all policies for individuals who perceived the policy to be ‘very effective’ compared to those who did not (p-value < 0.10) (Supplementary Material C). The largest average marginal effects were for banning all visitors (0.275, p-value<0.01) and for shutting down public transportation (0.254, pvalue< 0.01). The smallest average marginal effect was for banning large gatherings (0.083, pvalue< 0.01) (Supplementary Material F).

## 4. Discussion

We investigated the extent to which the likelihood of support for government outbreak response policies varies with outbreak severity using hypothetical vignettes. We also investigated how likelihood of support varies by health, income, age and perceived effectiveness of a policy. One of the main findings in this study was that the probability of support for any particular policy was primarily driven by outbreak characteristics. This is an important finding as (with the exception of Cook et al) previous studies did not incorporate outbreak severity when investigating public support for government response measures. We also found that the probabilities of support for all policies were greater for scenarios where the outbreak was more severe. We observed more variation in likelihood of support for various policies in cases of less severe outbreaks while likelihood of support was very high for all policies in scenarios with severe outbreak. These findings suggest that governments should be mindful that the public’s support for policies may change as an outbreak evolves. Consistent with our hypotheses, fatality rate was the most influential factor among outbreak attributes for most internal policies while global disease spread was the most influential factor for border control policies on average (based on the attribute levels used in this study). These findings suggest that individuals are less worried about the spread of the infection as long as the disease is not fatal. However, findings might have been differed if we used a much larger spread of the infection. In addition, unsurprisingly, the situation across the world became more important when it came to controlling the borders against visitors or residents coming back from other countries.

In less severe outbreak scenarios, the lowest support was for shutting down public transport (30.4%). This could be explained by the low car ownership rate of roughly 10% (as of 2018) in Singapore (compared to roughly 80% in the US and just slightly below 50% in Europe [24]) while its local public transport serves 7.5 million riders a day [25]. Hence it is clear why most participants would be reluctant to support shutting down public transport as it would greatly restrict day-to-day mobility within the city-state. This finding would be relevant to other large cities where public transport is the main mode of travel, such as Seoul [26] or New York City [27]. This finding suggests that if there are plans to shut down public transportation, the government can consider providing alternative modes of travel or subsidies for alternative services for those who work in essential services and need to commute.

Unsurprisingly, a lockdown of all citizens received the second lowest level of support (45.1%) due to the disruption it would cause to each individual’s daily life. In the case of a lockdown, those with jobs that do not allow the possibility of working from home may face a loss of income. Mandating every member of a household to remain at home is also known to cause increased prevalence of domestic and child abuse [28, 29], general adverse effects to mental health and low productivity [30]. None of the other aforementioned studies [6, 7, 9] explored support for a policy on lockdown of all residents. However, the lower public support on lockdown compared to other policies could potentially be generalizable to other countries as its effect on individuals’ lives would be similar in many countries. The public protests against lockdown seen in countries such as the US [31] and Germany [32] during COVID-19 pandemic provide evidence of lower levels of support for a lockdown.

Disallowing social gathering received the highest support among internal policies. This is likely because it was the least restrictive internal policy presented in our study. Overall, the lower levels of support for restrictive internal policies suggest that governments have to invest effort in designing targeted public health messages explaining to the public why these measures are necessary and how they can help with outbreak response efforts.

Border control policies received greater support than policies for internal restrictions for any given outbreak scenario. There might be several reasons for this. Firstly, border control affects daily life less than internal control policies for most individuals. Secondly, individuals could be lacking understanding on the effectiveness of different policies. Disallowing social gatherings was perceived to be very effective by only 39% of our sample, compared to the 67% to 76% who found border control policies to be very effective. However, previous literature has suggested that social distancing is considerably more effective at transmission control than restricting borders [33-35]. Lastly, the occurrence of a disease outbreak is often associated with higher levels of ethnocentrism and xenophobia [36, 37].

We found that ‘number of countries with rapidly increasing cases’ was one of the main determinants of the likelihood of support for restricting the entry of visitors or quarantine of residents returning from any country. This is understandable because more countries reporting rapidly increasing numbers indicates higher rates of transmission across the world. In this case, it can be seen that individuals preferred to adopt a more cautious approach, limiting exposure to all countries. This finding is also consistent with the response adopted by governments around the world during COVID-19. As COVID-19 cases were detected in more countries, governments implemented stricter border control measures.

We also found that the individual’s perception of a policy’s effectiveness as an outbreak control strategy was the most consistent predictor of support for all policies in our study. This is a noteworthy finding with important policy implications for governments. Our findings suggest that governments can focus on perceptions about the effectiveness of a policy to gain support from the public. Future studies can look into how public perceptions are influenced to increase public compliance to and support for a policy.

Among personal characteristics, less-healthy individuals (vs healthier individuals) were more likely to support all 4 border control policies, while individuals with lower-income (vs high income) were less likely to support 2 border control policies. We observed a health effect where less-healthy individuals had greater support for policies compared to healthier individuals in each income group, although the differences were significant only for the border control policies. On the other hand, we observed an income effect among both less-healthy and healthier individuals only for the lower-income group (in comparison to high-income group) for the 2 border control policies. These findings suggest that health concerns seem to outweigh income concerns during an outbreak for our sample. However, results may also be driven by lack of power. Future research should test whether these findings hold in other countries.

Age was found to be a significant predictor of the likelihood of support for most policies. Older individuals (vs younger individuals) were more likely to support most policies compared to those who were younger, with the exception of shutting down public transportation, which older individuals were less likely to support. The finding on shutting down transportation is not unexpected as 50.4% of car owners in Singapore are aged 30 to 39, which is higher than most age groups [38]. Hence younger individuals are less likely to be affected in the absence of public transport. Additionally, due to the stronger ideals of frugality among the older generation observed in many cultures [39, 40], older individuals are less inclined to spend money on private ride services, such as taxis. Overall, since older individuals were in general more likely to support response policies, public health messages should be tailored to appeal to younger ages.

The main limitation of our study is that some of our findings may not be generalizable beyond Singapore, and is perhaps least generalizable to countries where political ideology is vastly different, such as the US or France. Results may be most likely to generalize to similar Asian societies in developed nations such as Japan. Our study findings are also based on hypothetical vignettes and individuals’ preferences may be different during real outbreak situations. However, we minimize the effects of this limitation by implementing the study during the COVID-19 pandemic.

## 5. Conclusion

Our study is one of the first studies to investigate the likelihood of support for government policies in response to varying infectious disease outbreaks, and is the first study to investigate public support for very restrictive policies such as lockdown. Our findings showed that likelihood of support varied across government response policies; however, was generally higher for border control policies compared to internal policies. Individuals were willing to support even the most restrictive policies when the outbreak was severe. The fatality rate was the most important factor for internal policies while the degree of global spread was the most important for border control policies. Our findings suggest that campaigns to promote public support and compliance should be designed specifically to each policy and tailored to different segments of the population. They should also be adapted based on the evolving conditions in order to receive continued public support.

## Data Availability

Data used in this study is available from the corresponding author upon reasonable request.

## Acknowledgement

We would like to thank Dr Irene Teo for her feedback on the questionnaire.

## Declarations

### Data availability

Data used in this study is available from the corresponding author upon reasonable request.

### Code availability

Code used in this study is available from the corresponding author upon reasonable request.

### Authors’ contributions

All authors contributed to the writing and/or critical review of this manuscript.

